# Markovian Structures in Modelling COVID-19

**DOI:** 10.1101/2022.09.23.22280294

**Authors:** Elvis Han Cui, Weng Kee Wong

## Abstract

The aim of this paper is to model COVID-19 based on Markov chains. First, we introduce basic concepts of Markov chains with examples from different disciplines. Second, we use different types of Markov chains to model COVID-19, including confirmed cases, death and recovered cases and forecasting future confirmed cases. Third, we give conclusions based on these models and ideas for future work.

## 1 Introduction

COVID-19 has been a big threat in the U.S. since March, 2020 [1, 2]. Many people including us are affected by this unexpected disease. As statisticians, we would like to explore and predict the progress of COVID-19 from several perspectives. For example, how long will the current pandemic caused by COVID-19 last? How many confirmed cases will there be in summer? How different causes will affect the fatality rate later on?

This paper assumes the axioms of probability theory proposed by the Soviet mathematician A. N. Kolmogorov, and we study COVID-19 using a probabilistic framework. We regard all the information contains randomness. For instance, the daily number of confirmed cases is not deterministic and we believe is likely better represented by a curve that goes up and down in a probabilistic way. “Up and down” suggests some randomness in the numbers and the randomness comes from various uncontrolled sources and the inherent unpredictability of the disease. Consequently, it is a challenging statistical problem to model the disease progression of COVID-19, especially when countries adopt different strategies to control the spread of the disease. Our approach to study the disease is to use Markov chain models.

The next few subsections review basic concepts, advantages and limitations of Markov chains and common Markov chain models. Section 2 connects Markov chains with the SEIRD models commonly used in epidemiology. Each letter in SEIRD has a meaning with S for the number of susceptible, E for the number of exposed, I for the number of infected, R for the number of recovered (or immune) individuals and D for the death. In section 3, we describe a two person transmission model. In section 4, we extend the two person transmission model to a larger population. In section 5, we propose potential future work and conclude the paper.

### 1.1 Essentials of Markov chains and conditional independence

Intuitively, a Markov Chain (MC) represents a simplified independence relationship. Given the present state, the future is independent of the past. Mathematically, a Markov Chain is a collection of random variables {*X*_*t*_ : *t* ∈ ℝ^+^} where ℝ^+^ = [0,∞) and they satisfy the following relation, where a.s. here and elsewhere, stands for almost surely and *A* is a subset of the state space of *X*_*t*_.

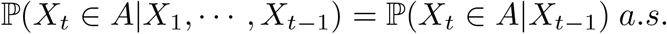

A Markov chain is called **homogenous** (HMC) if ℙ (*X*_*t*_ ∈ *A*| *X*_*t*−1_) does not depend on *t*. As an example, this means that *X*_*t*_|*X*_*t*−1_ has the same distribution as *X*_1_|*X*_0_. As a concrete example, consider a die-hard gambler deciding whether to continue to bet depends only on whether he has money or not, regardless of how many games he has played and the total number of games is interpreted as “time” here. In this case *A* is the event whether the gambler bets again at time t and *X*_*t*_ is the amount of money he/she has at time t-1. A Markov chain is called **non-homogenous** (NHMC) if ℙ(*X*_*t*_ *X*_*t*−1_) depends on *t*. More generally, we can define the Markovian structures on graphs, which is the theoretical foundation for graphical models. Formally, we say *X*_1_ and *X*_2_ are independent conditioned on {*X*_3_,*…, X*_*n*_} if for any sets *A*_1_ and *A*_2_:

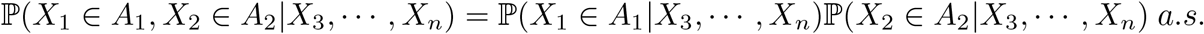

The conditional probability ℙ (*X*_*t*_|*X*_*t*−1_) is called the **transition kernel** of the Markov chain. For example, ℙ (*X*_*t*_ = *j*|*X*_*t*−1_ = *i*) = 0.5 means that the probability is 0.5 going from state *i* to state *j*. For more details and mathematical treatments on Markov chains, see [3].

### 1.2 Examples, advantages and limitations of Markov chains

Conditioning is one of the most important concepts in probability and Markov chain is the simplest and perhaps the most widely used probability model that exploits heavily on conditioning. Many interesting natural phenomenons can be simplified as a Markov chain. We give two examples below:

- the Bernoulli-Laplace model in physics describes the gas exchange in an airtight container. Imagine there are 2 containers with a wooden board blocking them. One container has k red balls (a kind of gas) and the other contains k blue balls (another kind of gas). Suppose a swap is made at random with 1 ball each from of the two urns, i.e., pick one from the red container and another one from the blue container, and then swap them. At the *t*^*th*^ swap, the distribution of balls in each container depends only on the number of balls at the (*t* − 1)^*th*^ swap and is independent of the previous (*t −* 2) states; see Figure 1.
- Brownian motion is derived as the limiting case of a Markov chain and describes the random motion of particles suspended in a fluid resulting from their collision with the fast-moving molecules in the fluid. Figure 2 is an example of Brownian motion on the plane with different colors representing different times. For example, red points correspond to movements from *t* = 0 to *t* = 200. Precisely, consider a particle starting at the point (0, 0). At each time, it has 4 possible movements: up, down, left and right and each movement has probability 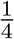. Knowing its location at time (*t*−1), the location at time *t* is independent of all previous locations. Such phenomenon describes a sense of “total randomness” in real life.

**Figure 1:**
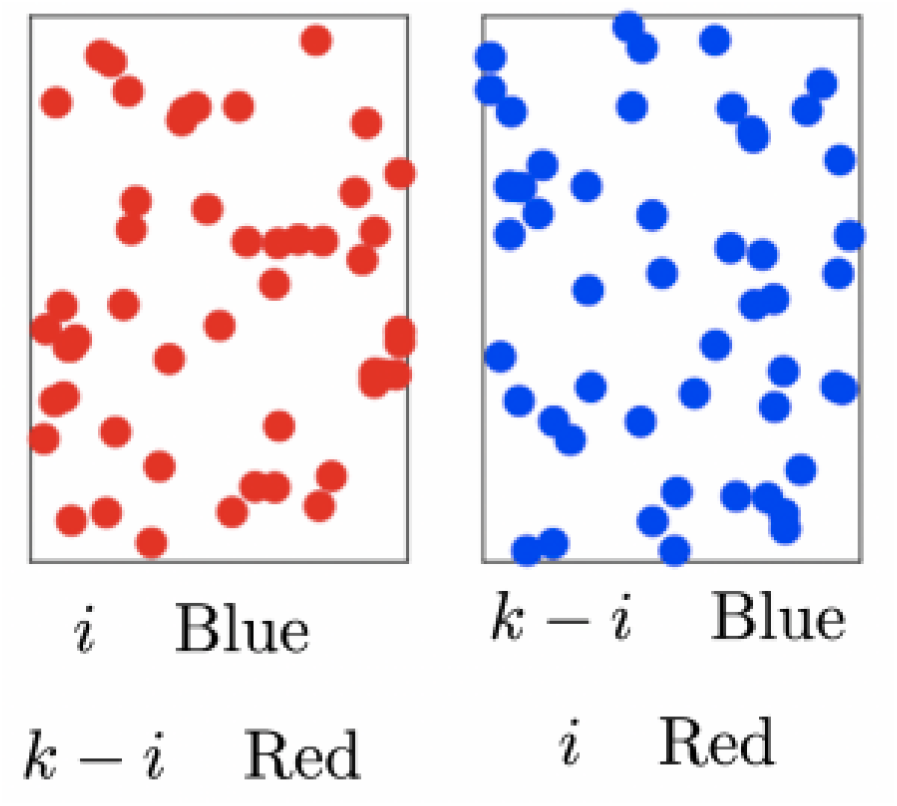
bernoulli-laplace model.

**Figure 2:**
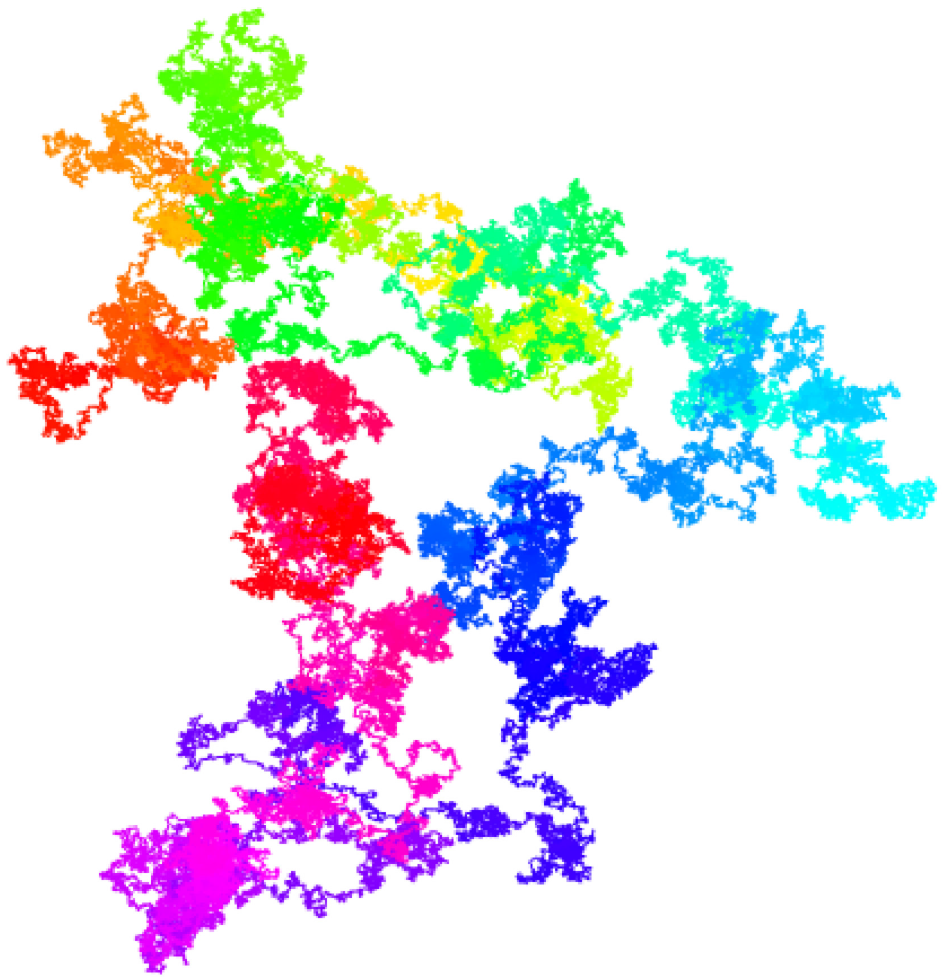
Brownian motion on the plane for t from 0 to 2000.

There are more phenomenons in real life that are simplified and studied using a Markov chain model. Markov chain is also applied to epidemiology and section 2 shows one of the models in epidemiology known as the SEIRD model is a realization of Markov chains; see section 4 for further details.

Despite the success and intriguing simplicity of Markov chains, they have several limitations for real applications

- The real world is complicated, and oftentimes a Markov chain model can only accommodate very few covariates. Consequently, MC is frequently unable to model adequately the process we observe in practice.
- Markov chains are based on the “conditional independence” proposed by Kolmogorov (Feller, 1971). In real life, existence of such a relation is not obvious and can be problematic to validate the assumption that conditional independence holds in practice.

### 1.3 Stochastic models for COVID-19

Our main interests are to predict future trends of the disease with accuracy and reliability. In our case, immediate questions of interest for our study are given the current COVID-19 data, what are the expected numbers of infected, suspectible, latent and the recovered subjects from COVID-19 in the future? More precisely, can we use the current data and use a model to predict the number of cases 3 months or even years from now? To do this, we use a model and postulate the stochastic nature of COVID-19. We could either explore this issue from a biological point of view and ask, for example, how COVID-19 attacks human tissues, and what kinds of cells are able to defend themselves from being attacked? Or, we can model the transmission of COVID-19. In this paper, we consider only the latter. The concept of transmission in epidemiology and public health can be abstracted as transition kernels in mathematics (Feller, 1971). Therefore, the question becomes how do we use Markov chains to model COVID-19. More generally, the research questions include the following:

1. Can we model the transmission of COVID-19 as a Markov chain?
2. What assumptions do we have to make?
3. Can we simulate the transition process of COVID-19 based on Markov chains?
4. How can we trust the results given by Markov chains?
5. Will there be more refined Markov structures or other stochastic structures in COVID-19?

The first three questions are about constructing models and identifying model assumptions. Question (4) is about advantages and drawbacks of Markov chains which we have discussed in the previous sections. The last question is more sophisticated and can be answered using graphical models. Some key assumptions will be stated in later sections and models will be built based on them. For the last question, another stochastic structure that we can explore is the spatial distribution of COVID-19. Understanding the spatial distribution allows us to answer question like what is the distribution of COVID-19 cases in the U.S.?

Other interesting questions can be asked. For instance, are there clusters in the locations of confirmed COVID-19 cases ? Some states have higher confirmed cases and others are lower. Is it due to human intervention (policy, state culture, etc.) or is it due to randomness ? Such hypotheses can be tested under the framework of Poisson random measure, which is a powerful tool for testing spatial homogeneity assumptions (Feller, 1968).

## 2 SEIRD model and Markov chains

Epidemiologists frequently use models to study disease development and progression over time. Some are complicated and require advanced statistical techniques. This section discusses common models for studying COVID-19, including the pros and cons, and their connections with Markov chains for each of the models [4, 5].

### 2.1 Basic concepts of SEIRD model

In epidemiology, SIRD models and its variations are applied to predict the future trend of COVID-19. A variation of SIR, which is more commonly used, is called SEIRD where the letter E additionally stands for exposed.

The SIERD model is defined by a system of differential equations:

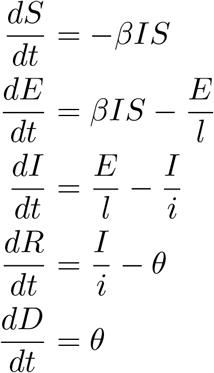

where *t* is time, *β* is the transmission rate of the disease, *l* is the latent period of the disease, *θ* is the death rate and i is the infectious period. These equations model a disease within a closed population, even though that population grows or shrinks birth and death process.

### 2.2 Connections between SEIRD model and Markov chains

The solution to the above differential equations, if it exists, is unique and deterministic. When the solution to the above equations exists, it is a collection of 5 functions, and each is a function of the time *t*. These differential equations are continuous operators and they are solved after they are discretized over the time space **t**, where **t** ranges over the natural number ℕ = {0, 1, 2, …}. For each time point *t*_0_, the state space (*M*, 𝕄) is (ℕ^5^, ℬ (ℕ^5^)) where 𝕄 = (ℕ^5^) stands for the collection of all subsets of ℕ^5^ and (*S, E, I, R, D*) is a 5-dimensional random vector taking values in ℕ^5^. Given all other model parameters (i.e., reproductive number, infectious rate, recover rate, etc.), the system of differential equations assume that the number of cases at the (*t* + 1)^*th*^ day is independent of the number of cases before (*t*−1)^*th*^ day. The only difference between a SEIRD model and a Markov chain is that the latter is random and the former is totally deterministic, where the transition kernel is a Dirac delta measure.

### 2.3 An numerical example based on COVID-19

To demonstrate the utility of the SEIR model, we provide an example with user-supplied initial values. The initial values are

- Suspected *S*_0_: 327200000. We assume the number of the suspected is the current population of the U.S. (https://www.worldometers.info/world-population/us-population/).
- Infected *I*_0_: 1. We assume there is only one individual that is infected at the beginning (*t* = 0).
- Recovered *R*_0_: 0. We assume no one is immune (recovered) at *t* = 0.
- Exposed *E*_0_: 10. We suppose there are 10 exposed individuals at *t* = 0.
- Death *D*_0_: 0. We assume no one is dead at *t* = 0.

Figure 3 shows the time trends of the numbers of death, recovered and infected individuals over time. It is generated by R 3.6.1 (https://www.r-project.org/). From Figure 3, we observed that the infected number increases at first and then declines. There is a time lag between the number of recovered cases and the number of cases of death.

**Figure 3:**
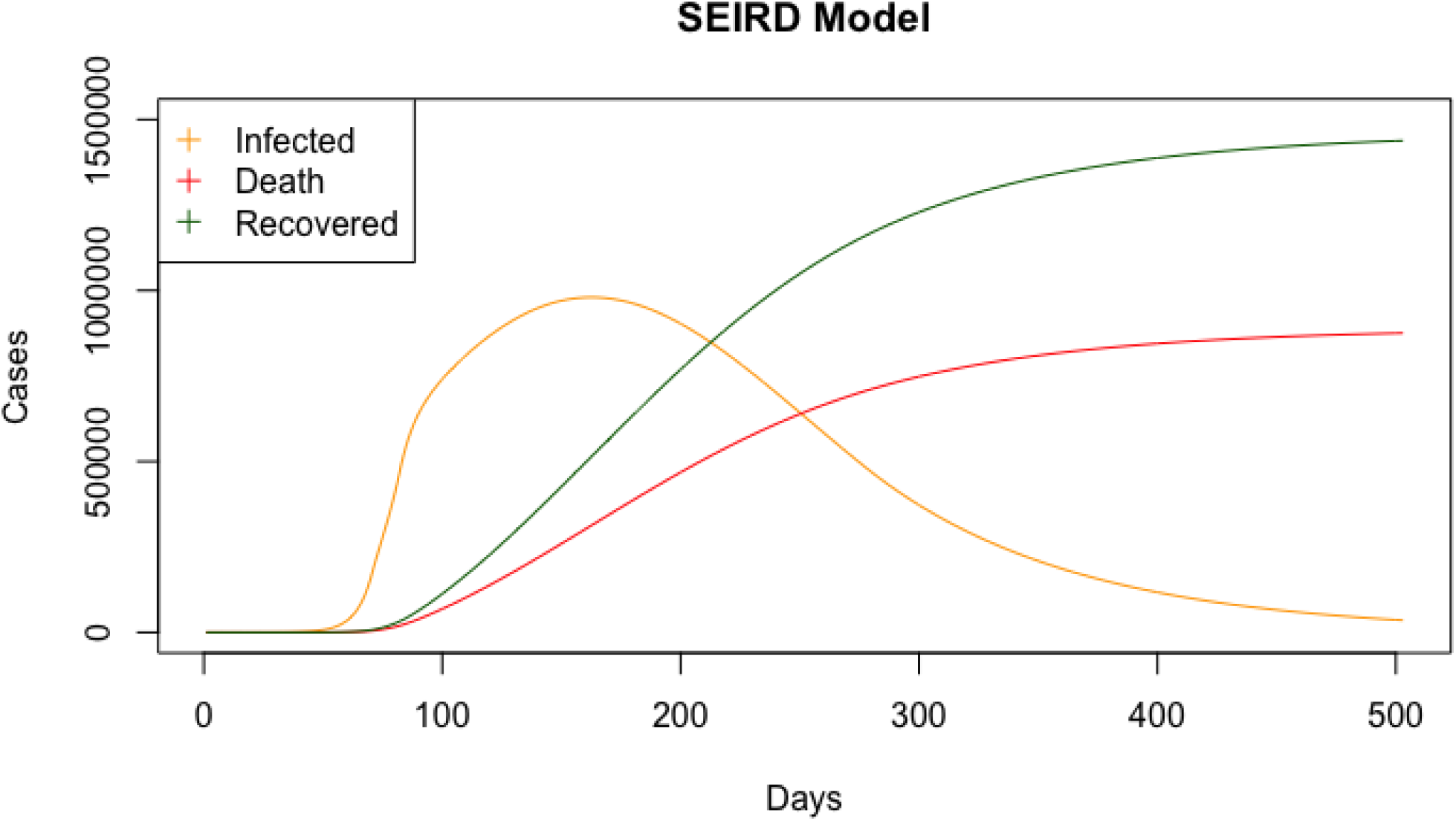
Predicted numbers from the SEIRD model for t from 0 to 500.

## 3 Two person transmission model

This section serves as prelude to the next section where we build more complicated models [6]. Suppose the state space of a person’s condition is {*H, C, D, I*}, where H, C, D, I stands for healthy, COVID-19 (infected), dead and immune (recovered) respectively. For a two person transmission model, there are 16 possible states in total: {HH,HC,HD,HI,CH,CC,CD,CI,DH,DC,DD,DI,IH,IC,ID,II}. A two person transmission model assumes that given current states of two people, the next states for the two people can be determined by a probability transition matrix. The matrix is 16 ×16 and its elements are the transition probabilities for a person to move from one state to another. Here is an example with the following assumptions for the two person transmission model to show how the transition kernel is constructed.

- If one of the two is already infected and the other is healthy, then the healthy one gets infected with probability 0.3 and the healthy one stays healthy with probability 0.7.
- For the infected one, regardless of the state the other has, he/she dies with probability 0.05 and recovers with probability 0.16 and stays infected with probability 0.79.
- If one has died, then he dies forever.
- If one gets immune, then he/she will never get infected again.
- Then a direct calculation shows the transition kernel is: For example, *p*_*HC,HC*_ = 0.55 (element in (2,2)) means if the first person is healthy and the other is infected, then the probability is 0.7 ×0.79 ≈0.55 that the first stays healthy and the other stays infected. As another example, if *p*_*HC,CC*_ = 0.24 (element in (2,6)), this means that if the first person is healthy and the other is infected, then the probability is 0.3 *×* 0.79 *≈* 0.24 that both of them are infected.

### 3.1 Long-term behavior

We are interested in the long term behavior of the transition matrix. Given a initial state (say, HC, one is healthy and the other is infected), what are the states for the 2 people in the long run? Simulation results are below.

The left figure is the graphical representation of the 16 ×16 transition matrix. The darker the color, the higher is the probability of attaining that particular state (column indexes) for a patient in the current state (row indexes). Diagonal elements have higher probabilities which means the system prefers current state than other states. For example, the top left element means the probability is 1 from state *HH* to state *HH* and is 0 from *HH* to any other 15 states. The element 𝒫_2,2_ means the probability from state *HC* to *HC* is 0.55.

The right figure shows the long term behavior of the transition matrix. If the transition matrix is 𝒫, then the long term behavior matrix is 𝒫^*∞*^ = lim_*n*→∞_ 𝒫^*n*^. Convergence and other theoretical properties are guaranteed by Perron-Frobenius theorem (Dabrowska, 2020). The elements of the long term matrix 𝒫^*∞*^ are obtained by multiplying 𝒫 repeatedly without end. For our example, the long term matrix is:

For example, the element 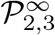 means starting from the initial state *HC*, on average, the probability is 0.08 that one person stays healthy and the other is dead. “On average” corresponds to “long-term”. Except for absorbing states (states with transition probability 1), state *II* has a significantly larger probability among all initial 16 states, suggesting that herd immune is mostly possible in this model.

## 4 Non-homogeneous Markov chain model

In this section, a non-homogeneous Markov chain (NHMC) model is applied to simulate the transmission of COVID-19 within a region under certain conditions. This model is inspired by Conway’s game of life (https://en.wikipedia.org/wiki/Conway) and the work in [7].

This model requires several assumptions:

1. The whole region is a *p × p* grid, i.e., each cell represents a person (or a group of people).
2. Each has 5 states: healthy (H), latent (L), COVID-19 (C) (or infected), dead (D) and immune (I) (or recovered). Note that “latent” stands for a person is infected but no on-site symptom is observed and COVID-19 means the symptom is observed.
3. Each is essentially fixed at his own cell (i.e. as time goes by, the individual doesn’t move to other cells).
4. After each time unit (a week, a month, etc.), the individual changes state from *i* to *j* where *i, j* ∈ *{H, L, C, D, I}*.
5. The transition probability of a person is determined by the neighbors surrounding him (i.e., first order relation).

### 4.1 Model set-up

In this section, we introduce basic elements of the model: initial state, transition kernel and one-step update. We use the JHU dataset (https://github.com/CSSEGISandData/COVID-19) to give point estimates of the transition kernel. The JHU dataset contains information on state-level confirmed, recovered and dead cases in the U.S. For more details on how the data is collected and more visualizations on the dataset, see https://coronavirus.jhu.edu/us-map.

#### 4.1.1 Initial state

In this subsection, we generate the initial state of the model randomly. We suppose the grid is 88 ×88, which means that there are 7744 individuals. According to current infection rate in the US (667,000 out of 390,000,000), 0.1% people are in state *C* (have COVID19) and 0.3% people are in state *L* (who are infected and no on-site symptoms is observed). The choice of 0.3% is based on the personal communication with Dr. Ramirez in Biostat244. Others are in state *H*. We generate the initial states of individuals independently: with probability 0.1%, the individual has detectable symptoms, with probability 0.3%, the individual is infected but has no detectable symptoms and with probability 99.6%, the individual is healthy. Figure 7 displays the initial state of the non-homogoneous Markov chain, where color purple stands for *H*, light green stands for *L* and yellow stands for *C*: From Figure 7, we observe there are 9 people in state *C* (yellow), 18 people in state *L* (light green) and 7717 people in state *H* (purple). People in state *L* and *C* are uniformly distributed across the grid and we are interested in what the grid will be after a long term.

**Figure 4:**
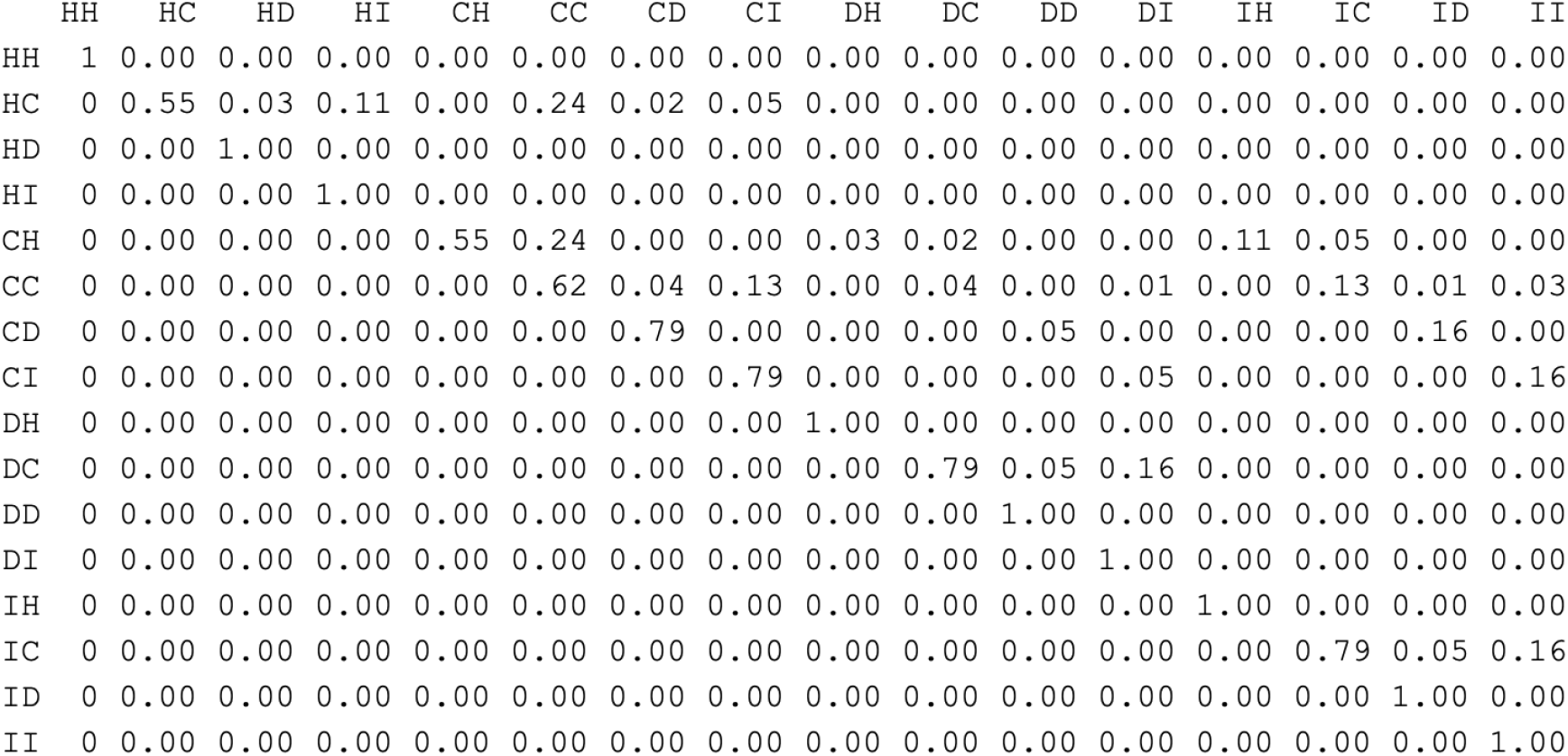
Transition kernel for the two person transmission model.

**Figure 5:**
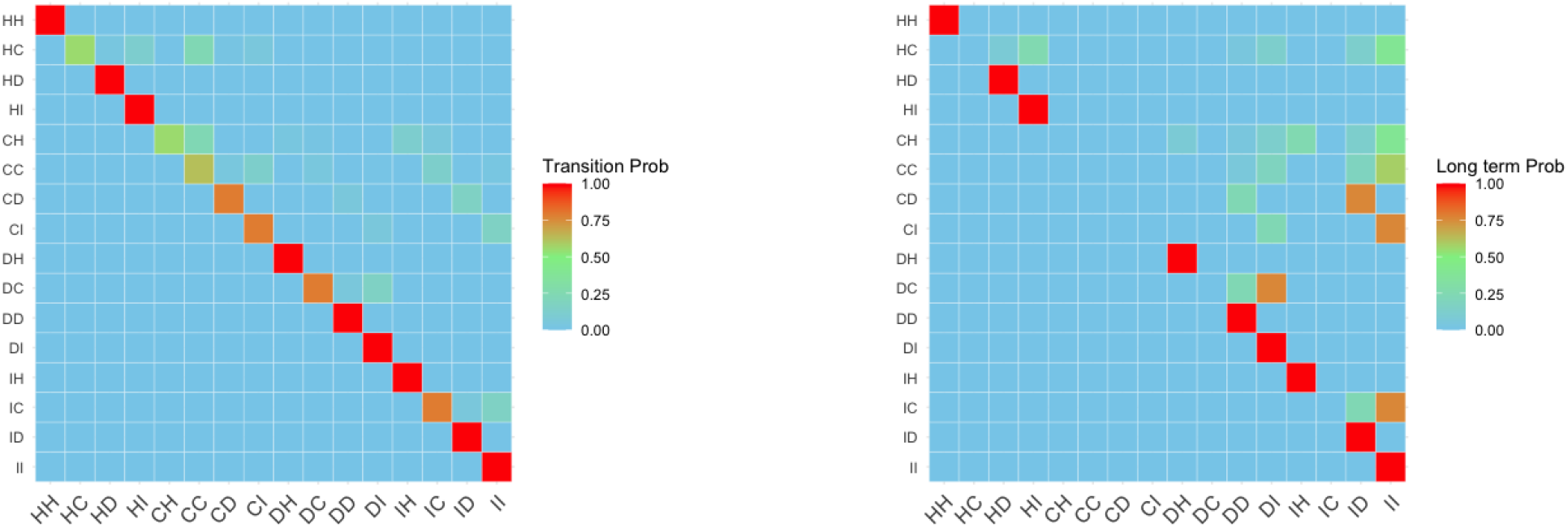
Transition matrix (left) and long term transition matrix (right).

**Figure 6:**
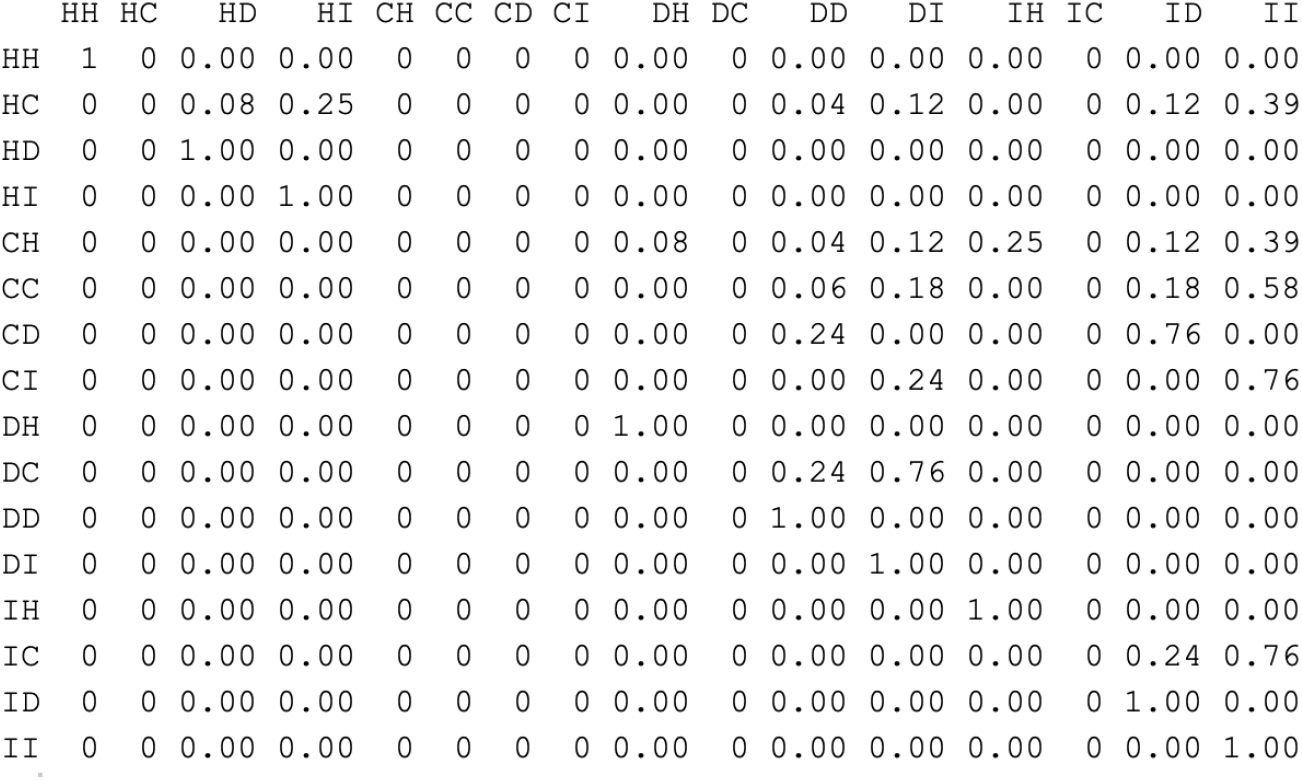
Long term transition matrix 𝒫^∞^ for the two person transmission model.

**Figure 7:**
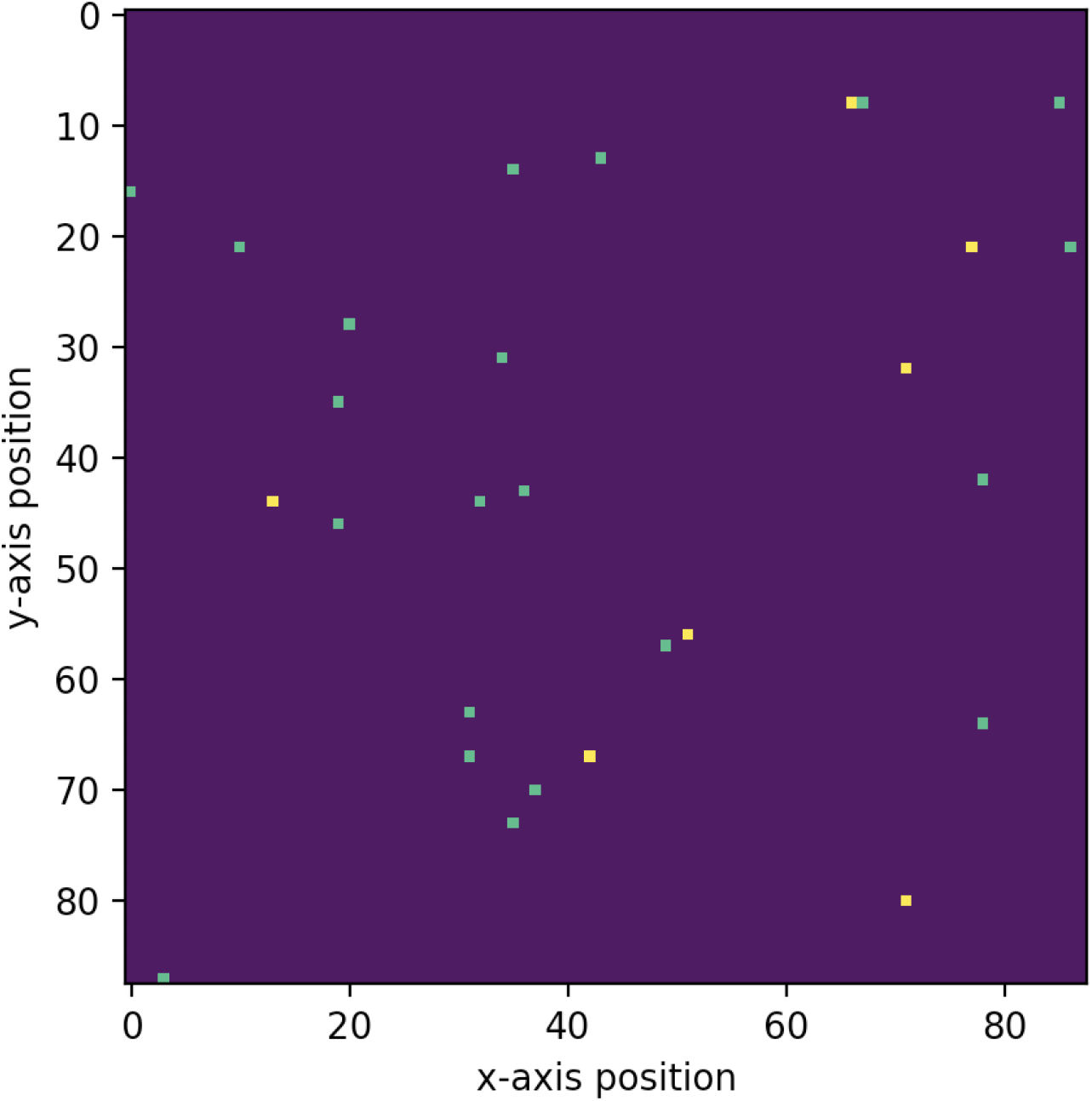
Initial state of NHMC at *t* = 0.

#### 4.1.2 Transition kernel (transition probability matrix)

To determine the transition kernel of our model, we have to borrow existing results for some key probabilities. For example,

1. **Fatality rate**: By April 16, there are 27,697 confirmed cases in California and 956 deaths due to COVID-19. If a person has detectable symptoms, the probability of dying can be estimated as 956 / 27697 which is 0.034. Although the rate for individual is time-dependent, we assume it is homogeneous.
2. **Recover rate**: This corresponds to the transition probability from state *C* to state *I*. There are 1,473 recovered cases and 27,697 confirmed cases in California, so the recover rate is 0.053. We also assume the rate is homogeneous, i.e., independent of age, infected time and other factors. Similar to previous section, we assume *p*_*II*_ = 1. If a person recovers, then he never gets infected again. Using the language of Markov chain, state *I* is called **absorbing**.
3. **Infection rate**: This is similar to the *R*_0_ value in epidemiology. In this model, we calculated the state of 8 neighbors surrounding a person (3 and 5 if he is in the corner and border), summing up the number of state *C* (COVID) and *L* (latent). The probability of from healthy to infected is an increasing function of that sum. We set the base infectious rate to be 0.8, that is, if one healthy person has only one infected neighbor (on-site symptom or latent), then in the next time unit, the probability of him getting infected is 1 −0.8 = 0.2 (from state *H* to state *L*). If he is surrounded by *k* infected people, the probability is 1 −0.8^*k*^ and 0 if no infected neighbor.

Given the information of neighbors, the transition kernel for an individual is a 5 *×* 5 matrix with the following form:

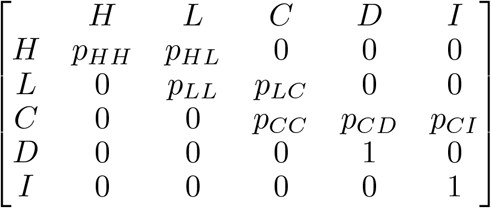

Importantly, this matrix is different from those in Markov chains that are homogeneous in time because we are conditioning on the state of one’s neighbors.

#### 4.1.3 One-step update

Before each update, we need information on the locations of infected and latent people to calculate the transition kernel, and the computation steps are:

1. Scan current states for all individuals in the grid. It requires *p*^2^ operations (in our case, *p*^2^ = 88^2^ = 7744 individuals).
2. For each individual, determine the information of the 8 neighbors (whether they are in state *L* or *C*). This process requires 8 operations for each individual.
3. Derive the transition kernel for each individual based on current state and this operation requires *p*^2^ operations.
4. For each individual associated with a unique transition kernel, we determine the next state through sampling. It requires 5 operations for each state and the total number of operations is 5*p*^2^.

Putting the above operations all together, the time complexity for each update is approximately 15*p*^2^, or *O*(*p*^2^) (**time complexity** quantifies the amount of operations required in each update).

### 4.2 Simulation study

To better understand how a non-homogeneous Markov chain (NHMC) mimics the transmission of COVID-19, we conduct the following simulation study. The codes are written in Python 3.7.1 and is free online (https://github.com/ElvisCuiHan/Lecture-notes-on-Statistical-Learning/blob/master/MC_COVID.py). We ran 50 iterations (*t* = 1, …, 50) because when *t* is more than 50, the chain shows no significant changes of states, suggesting the chain has become stable.

#### 4.2.1 Long term behavior

The long term behavior of a Markov chain implies how the system will be after a long period of time. Figure 8 shows how the system enters a stable distribution after approximately 50 time units. Although the number of people in state *C* (people with detectable symptoms) is only a half of the number of infected people (in state *C* or *L*), both of them reach their peak at *t* = 20. As *t* increases, most people recovered and others are dead. When *t* is greater than 40, the chain enters a **steady state**, no significant fluctuation in confirmed cases, recovered or dead. There are more than 6000 people (out of 7744) get recovered and approximately 1000 of them are dead. The graph is similar to those from the SEIRD models and it suggests a faster outbreak of COVID-19.

**Figure 8:**
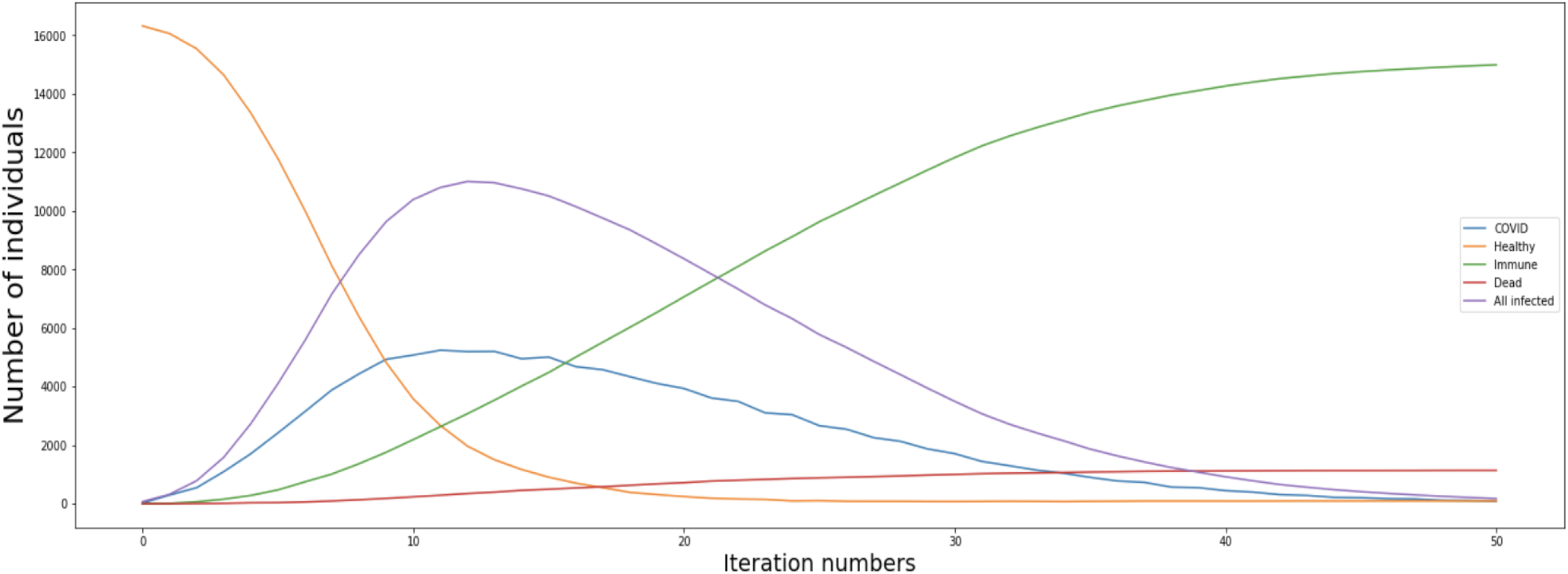
Long term behavior of NHMC for *t* from 0 to 50.

#### 4.2.2 Dynamic results

The long term behavior does not tell how individual is affected during the transmission of COVID-19. This subsection illustrates how COVID-19 spreads from a microscopic point of view, and Figure 9 gives details about the intermediate period of the whole system infection process with different colors representing different states:

- Purple: H (healthy)
- Light green: L (latent or without detectable symptoms)
- Yellow: C (COVID or infected)
- Green: I (immune or recovered)
- Blue: D (dead)

**Figure 9:**
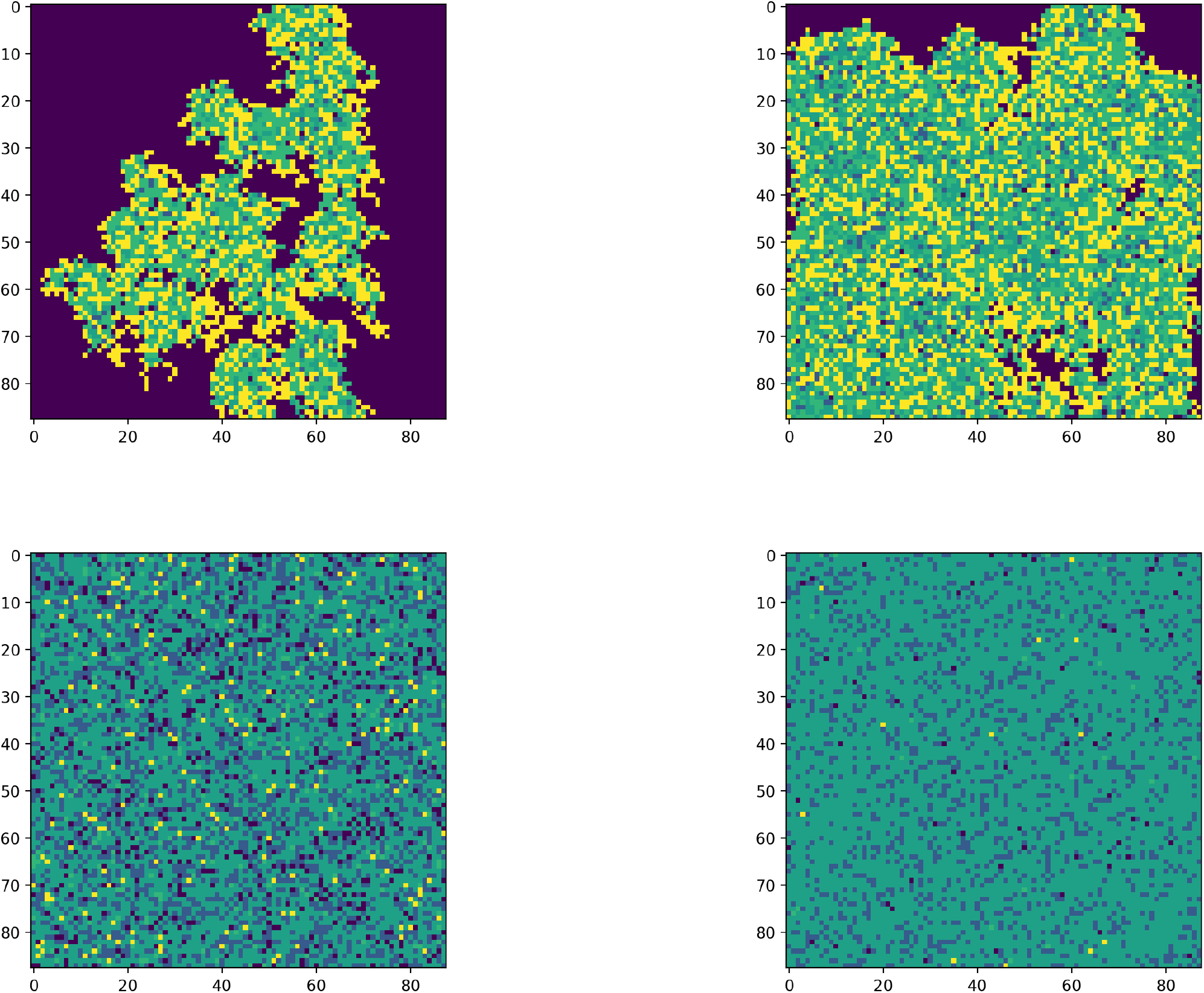
Early, middle, late and stationary states of COVID-19 as modeled by a NHMC.

The x-axis and y-axis represent the location of individuals. For example, the (88, 0) point in the top-left panel means the individual was healthy at the early stage. In the top-right panel, the (88, 0) point became yellow, suggesting that the individual was infected at the middle stage. The individual got immune according to the bottom two panels. Different plots in Figure 9 correspond to different periods and they give us valuable details on how exactly COVID-19 spreads out. For example,

- Top-left (*t* ≤10): The early stage of the model. Comparing it with the initial state, more people are infected and many of them don’t have detectable symptoms (in state *L*). Therefore, the detectable number is still small. Further, individuals are likely unable to realize that many of them are already infected.
- Top-right (10 *< t* ≤20): The middle stage of the model. It is the outbreak period of COVID-19 where many individuals are infected (in state *C* or *L*) and almost half of them have detectable symptoms. The number of death is increasing and we speculate that individuals are getting panic about the disease.
- Bottom-left (20 *< t* ≤35): The late stage of the model. The number of death continues increasing. More and more individuals get recovered, which is a signal that COVID-19 may disappear (or become less severe) sooner or later.
- Bottom-right (35 *< t*): The stationary stage of the model. The recovered, dead and infected rates become steady and there were no more significant fluctuations. COVID-19 seems to have almost disappeared after about 1000 lives were lost to the disease.

The model suggests that the number of patients with detectable symptoms is not reliable (top-left), the outbreak of COVID-19 is fast (top-right), a huge number of death is inevitable (bottom-left) and COVID-19 will disappear at some point (bottom-right). Compared to the results from the SEIRD models, non-homogeneous Markov chain model gives similar results except for the shorter outbreak period.

## 5 Future work

The previous sections suggest Markov chain model are useful to model a dynamic system such as in the disease progression and epidemiology. As pointed out in section 1, they are not without several drawbacks. For example, it only takes advantage of the information at the (*t*−1)^*th*^ state, regardless of the previous (*t*−2) states. One alternative is a polynomial regression where we use all information from *t* = 0 to the current state.

### 5.1 Non-Markov structure: polynomial regression models

A polynomial regression model assumes the confirmed cases of COVID-19 is a polynomial function of time. After we choose the degree of the polynomial to be fitted, confidence intervals can be constructed at selected time points. For example, if we use COVID-19 data for Los Angeles (LA) and the selected degree of the polynomial is 4, our codes (see https://github.com/ElvisCuiHan/Lecture-notes-on-Statistical-Learning) provide the following fit:

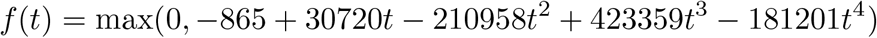

A 80% confidence band is given. The polynomial model is an example of **curve fitting model** described by [8]. Though it uses all information from the past, the variance of the fitted values can be high. As the degree increases, we are fitting noises instead of the data itself. Figure 10 shows the fit, along with a 80% confidence band for the fitted curve for the Los Angeles data from Jan, 22, 2020 (*t* = 0) to June, 19, 2020 (*t* = 1).

**Figure 10:**
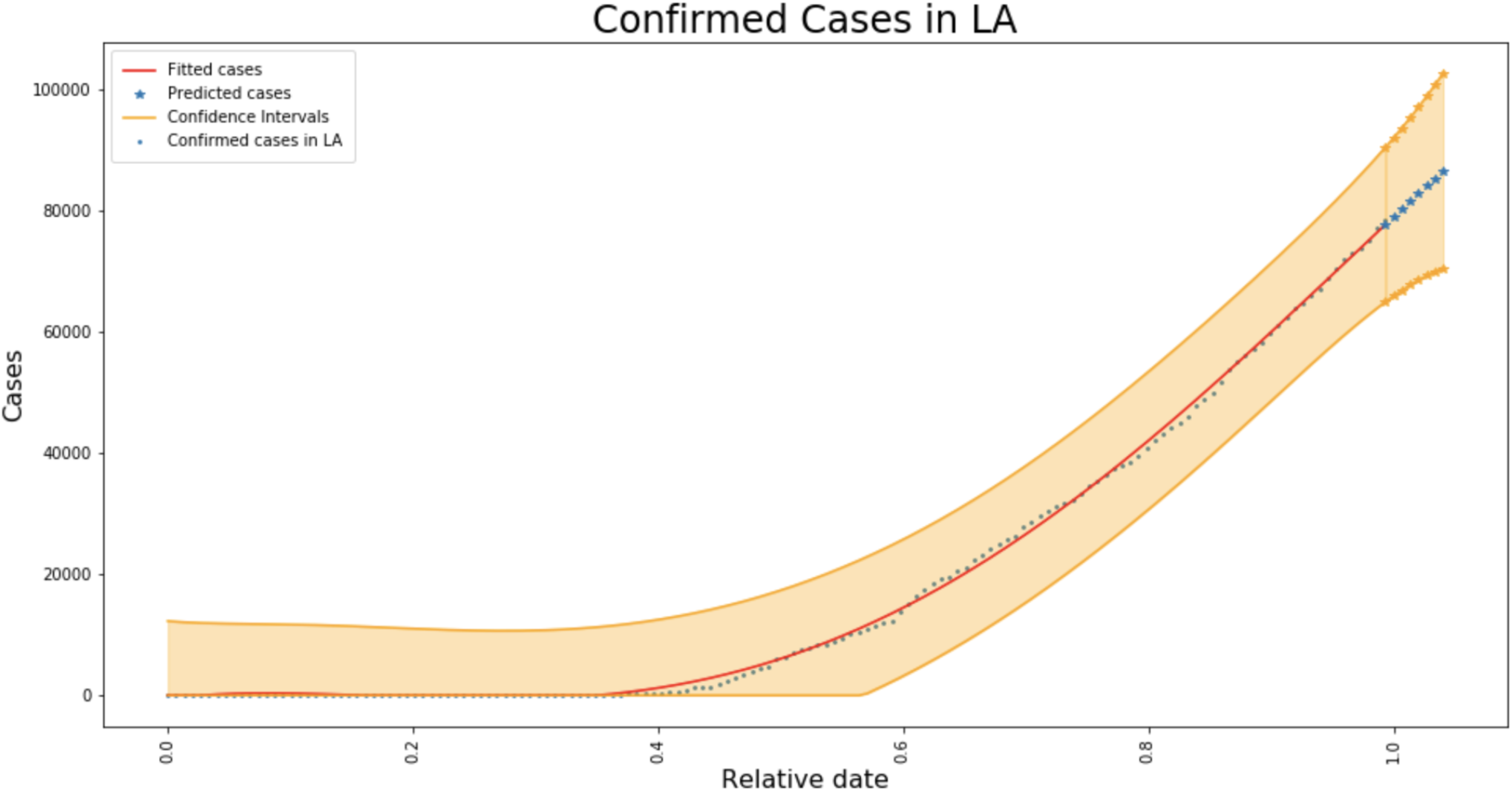
Polynomial regression for the confirmed cases in California.

### 5.2 Spatial distribution

In the non-homogenous Markov chain model, we assume the world is a *pI*× *p* grid which is not realistic. Two natural questions are:

- What is the spatial distribution of COVID-19 in the U.S. (or California)?
- Are the cases of COVID-19 randomly distributed?

Such questions require more detailed individual-level information on patients. They include history of their disease, where they have been to and where they live. The answers to these questions are difficult to collect which means that we have to make more assumptions to make our models more plausible.

### 5.3 Continuous time models

All previous models (SEIRD, two person transmission, non-homogeneous Markov chain, polynomial regression) are discrete time models. Though SEIRD is defined by a system of differential equations over a continuous time space, we have to discretize them to do simulations. This means that there is loss of information and it is more natural to construct a continuous time model. For example, instead of updating confirmed cases day-by-day, we could collect real-time data. This is very useful because this means that once a person has suspect symptoms, we can update the individual disease status instantaneously. Similar to previous subsections, data of this type is difficult to obtain. Our options are either to make more assumptions or perform another simulation study based on continuous Markov chains before data collection.

In conclusion, under the assumption of conditional independence, Markov chains are useful to simulate the dynamic process of the transmission of COVID-19 and they allow us to observe the process from a microscopic point of view. Despite of its convenience and usefulness, there are several additional works can be done such as extending the discrete time model to continuous time, exploring the spatial distribution of COVID-19, etc.

## Data Availability

All data produced in the present study are available upon reasonable request to the authors.

